# Plasma protein and tumor tissue gene expression analyses in ovarian cancer reveals differentially co-regulated clusters between benign and malignant conditions

**DOI:** 10.64898/2025.12.15.25342255

**Authors:** Mikaela Moskov, Julia Hedlund Lindberg, Ulf Gyllensten, Stefan Enroth

**Affiliations:** Department of Immunology, Genetics, and Pathology, Uppsala University; Biomedical Center, SciLifeLab Uppsala, Uppsala University, SE-75108 Uppsala, Sweden

## Abstract

Ovarian cancer is the deadliest of gynecological cancers and surgery is often necessary for a final diagnosis. Benign cases could be managed more conservatively, avoiding the risks and complications associated with surgery, if accurate diagnostic biomarkers existed. Underlying differences between circulating protein biomarkers and tumor gene expression also restricts interpretation and prioritization of potential biomarkers for diagnosis and potential drug targets. Here, high-throughput affinity plasma proteomics data encompassing over 5400 proteins in plasma from 404 women from two independent Swedish cohorts were analyzed alone and combined with total RNA sequencing in corresponding benign and malignant tumor tissue. A subset of 191 proteins previously identified as differentially expressed between benign and malignant conditions were used to perform correlation analyses, revealing similar patterns between groups but much stronger signals in malignant cases. Comparison with known protein interactions from the STRING database revealed a highly interconnected network consisting of 154 proteins in plasma. Differential correlation analysis (DCA) was performed on the full set of 5414 proteins and for their corresponding tumor RNA expression. DCA identified 31 plasma proteins with significant differential correlations (adjusted p < 0.05, ΔR > 0.5) and 759 tumor transcript pairs with significantly differentially correlating RNA expression. Distinct protein-protein correlation patterns in plasma were discovered and validated with notable differences between benign and malignant tumors. In general, these patterns were distinct from those detected on gene expression level in tumor tissue. In conclusion, our findings reveal clear differences in plasma protein co-regulation, with distinct correlation patterns between malignant and benign cases. The differences between results obtained in tumor transcriptomics and plasma proteomics results from the same patients warrants further studies into the tumor microenvironment to understand the function of promising protein biomarker candidates and the potential of these as future drug targets.

## Introduction

Ovarian cancer is the 8^th^ most common cancer amongst women worldwide, with a global mortality rate of over 200 000 women in 2022^*1*^. Although it is only the 3^rd^ most common gynecological cancer in the Western world, it remains the deadliest. In many countries, the 5-year survival rate of ovarian cancer is below 50%^*1*^, the main reason being that it is often diagnosed late and thus at a more advanced stage. If the cancer is discovered early, in stage 1, the 5-year survival rate is around 90%, while women with stage IV ovarian cancer has a survival rate less than 30%^*2,3*^. There is currently no reliable way to diagnose ovarian cancer in symptomatic women without surgery, nor is there a way to screen for it^*4*^. The biomarkers presently used, mainly cancer antigen 125 (CA-125, also known as mucin-16/MUC16), sometimes in combination with human epididymis protein 4 (HE4 / also known as WFDC2), are not sensitive enough to identify all cancers nor specific enough to clear benign cases^*5*^. CA-125 has a high sensitivity for late-stage cancers, but has low sensitivity for early-stage cancer^*6*^. It also has a high rate of false positive indications in many benign gynecological conditions such as infections, pregnancies, or endometriosis^*6*^. HE4 has higher specificity and is best used to separate benign from malignant cases but not for general population screening^*7*^, and also often elevated in other conditions such as endometrial cancer ^*8*^. Imaging techniques such as transvaginal ultrasound can achieve a very high accuracy in symptomatic women separating benign from malignant conditions^*9*^, but often require referral to tertiary centers and the accuracy rely on access to highly experienced practitioners^*7,9*^. For these reasons, symptomatic women must often go through surgery to get their final diagnosis. There are indications that benign tumors with no or even mild additional symptoms could be handled conservatively, avoiding the risk of long-term complications associated with adnexal surgeries^*10*^. Biomarkers for early detection and clear separation between benign and malignant tumors are needed to both improve overall survival, and the quality of life for affected women. Recent development in high-throughput proteomics, especially using high-sensitive affinity-based methods, have identified several plasma protein biomarker candidates for ovarian cancer^*11*^.

We have recently analyzed over 5400 plasma proteins and identified 191 potential biomarkers separating benign from malignant ovarian tumors^*12*^. These 191 candidates were validated using an independent cohort where we also characterized gene expression in tumor tissue from the same set of patients. Across both benign and malignant diagnoses, we found that for 11% of the 191 plasma biomarkers, protein expression correlated with the tumor gene expression^*12*^. In addition, we found several instances where plasma biomarkers showed no correlation with its corresponding gene expression, but where they were part of a larger protein-protein correlation network where at least one other member was correlated with its corresponding gene expression. This observation restricts the immediate interpretation of strong circulating biomarkers in relation to for instance their possibility to be a potential future drug-target, since differences in protein abundances could be related to down-stream regulation in the tumor itself or in the tumor microenvironment. Additional studies are therefore needed to further characterize regulatory networks in patient material containing both benign and malignant diagnoses rather than e.g. cell lines which do not replicate the surrounding environment. Here, we have further investigated the correlation patterns of protein and gene expression and, specifically, the differences in these correlation patterns between benign and malignant tumors.

## Experimental Procedures

### Experimental Design and Statistical Rationale

The samples and data used in this study is the same as previously described in Moskov *et al*^*12*^. In brief, a total of 404 blood plasma samples were collected from two independent cohorts from different geographical locations in Sweden. The samples were obtained from women who, after suspicion of ovarian cancer, were surgically diagnosed with either malignant (case) or benign (control) tumors. The plasma proteome of both cohorts was characterized simultaneously using high-throughput PEA, quantifying a total of 5420 proteins which were then further analyzed. Out of these proteins, 5414 were kept after removing replicates and those with missing values. A total of 191 proteins with significantly different protein expression between case and control were previously identified^5^ and these were used for the correlation and interaction analyses. The study was approved by the Swedish Ethical Review Authority (2023-05515-02, extension of Regional Ethics Committee in Uppsala Dnr:2016/145) and Göteborg (Dnr: 201-15). Informed written consent was obtained from all participants following the guidelines of the Declaration of Helsinki.

### Statistical analysis

The computational analyses were performed using R version 4.5.0^*13*^. Unless otherwise stated, p-values were adjusted for multiple testing using Holm’s method, and statistical significance was defined as adjusted p < 0.05. Networks were visualized using the igraph package^14,15^, with node sizes scaled between 3 and 30 based on connectivity and layouts optimized using the Fruchterman–Reingold algorithm. When multiple protein names were associated with the same UniProt ID, only the first was displayed. All plots were generated in R, and figures were finalized using Adobe Illustrator.

### Correlation analysis

Plasma protein correlations were calculated separately for the discovery and replication cohorts, as well as across all subgroups, using the 191 previously identified significant proteins. Pairwise correlations were computed with the corr.test() function from the psych package (version 2.5.3)^*16*^ using the Spearman method and Holm’s adjustment method. Correlation matrices were compared between cohorts. The heatmaps were produced using the heatmap.2() function of the gplots package (version 3.2.0)^*17*^ with custom inclusion of an additional line for the significant correlations.

### Protein-protein interactions

Protein interactions were extracted from the STRING database with the STRINGdb package (version 2.20.0)^18^ using version 12.0, species 9606 (human), a score threshold of 400, and a full network type. The 191 significant proteins were analyzed by mapping the UniProt and protein name to the database by the $map function of STRINGdb, and a weighted network was created using igraph, with unconnected proteins removed. Cluster communities were identified using the Leiden clustering algorithm based on modularity of the cluster_leiden() function of igraph, with interaction scores as weights.

### Differential correlation analysis

The DCAs were performed using the DGCA package (version 1.0.3)^*19*^. Pairwise Spearman correlations were computed using the ddcorAll() function with Holm’s adjustment for multiple testing. The results were filtered based on significance (p < 0.05) and correlation difference (ΔR > 0.5). For the proteomics data, the analyses were done separately for each cohort, and the discovery results were used as basis for filtering and visualization. Unweighted networks were constructed using the igraph package. As before, clusters were identified using the Leiden algorithm, but here without using weights.

## Results

### Plasma proteomics and gene expression data

The data used in this study has been previously published^*12*^. The proteome data includes measurements of 5416 unique plasma proteins using the proximity extension assay (PEA) and the Olink Explore HT panel, from 404 women diagnosed with either benign (control) or malignant (case) ovarian tumors. The PEA reports protein concentration in normalized protein expression (NPX), which is on a log2-scale. After quality control 5414 proteins remained. The samples are from two clinical cohorts from geographically different locations in Sweden. One cohort was used as a discovery cohort, and the other as validation cohort. The discovery cohort consisted of 171 individuals (86 controls and 85 cases) from Göteborg, and the validation cohort 233 individuals (81 controls and 152 cases) from the U-CAN biobank^20^ in Uppsala. The cohorts were matched with respect to age at diagnosis, where the average ages in discovery and replication were 60.3 and 61.5 years, respectively (p = 0.71, two-sided Wilcoxon ranked test). Total RNA-sequencing data from ovarian tumors, expressed as transcripts per million (TPM), was available for over 60 000 individual transcripts of 110 individuals (9 controls, 101 cases) in the replication cohort. Additional details on the cohorts and data generation can be found in Moskov *et al*.^*12*^.

### Protein-protein correlations reveal distinct patterns with major differences between benign and malignant cases

To investigate the differences in the correlations between plasma proteins for benign and malignant cases, pair-wise correlations for the previously identified^*5*^ 191 biomarkers candidates were calculated. First, all observations from the discovery cohort were used, with controls and cases combined, and then, controls and cases were studied separately (Supplementary Data 1).

From these analyses, distinct correlation patterns could be identified (Figure 1a-c), where the primary branches of the dendrogram in Figure 1a divided the correlations into two clusters, one smaller cluster with mainly negative correlations and one larger cluster with mostly positive correlations. This division can also be seen in the results from analyzing the control and case correlations separately (Figure 1b-c), where the results were ordered according to Figure 1a. In the combined analyses of both cases and controls (N=171), a total of 14226 pairs (78.4%, 14226/18145. 191 ^*^ 190 / 2, all unique protein-protein pairs, no self-correlation) were found to be significantly correlated (*q* < 0.05, adjusted for multiple hypothesis testing with Holm’s method (Figure 1d). Among these, 98.2% of the correlations were positive and 1.8% were negative. When analyzing the controls (N=86) separately, 12.1% of correlations were significant (Figure 1e). Out of these, 2146 (98.1%) of correlations were also significant in the combined analysis, with 100% of the overlapping correlations being positive. For the separate analysis of the cases (N=85), 46.2% of all pairs were found to be significantly correlated, out of which 8380 (99.9%) were also significant in the combined analysis, 99.5% were positive, and 0.5% were negative (Figure 1f). For a comprehensive summary of the number of significant correlations for each group and their distribution between positive and negative correlations, see Table 1.

**Table 1.**
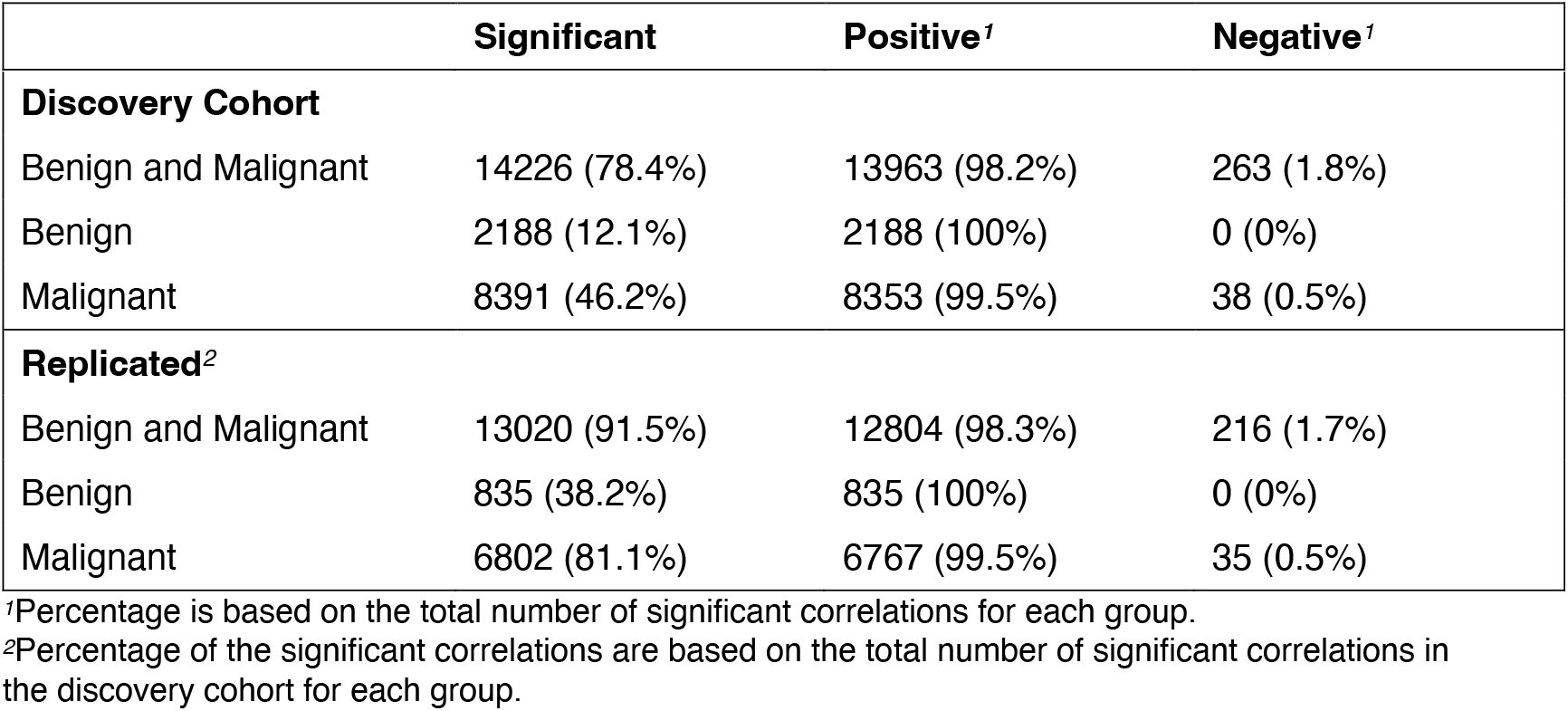
Number of significant correlations in the discovery cohort out of all 18145 possible protein-protein pairs (191 ^*^ 190/2, all unique protein-protein pairs, no self-correlation), the replicated significant correlations between cohorts, and the distribution between positive and negative correlations.

| Significant | Positive<sup>1</sup> | Negative<sup>1</sup> |
| --- | --- | --- |

| **Discovery Cohort** |  |  |  |
| --- | --- | --- | --- |
| Benign and Malignant | 14226 (78.4%) | 13963 (98.2%) | 263 (1.8%) |
| Benign | 2188 (12.1%) | 2188 (100%) | 0 (0%) |
| Malignant | 8391 (46.2%) | 8353 (99.5%) | 38 (0.5%) |
| **Replicated<sup>2</sup>** |  |  |  |
| Benign and Malignant | 13020 (91.5%) | 12804 (98.3%) | 216 (1.7%) |
| Benign | 835 (38.2%) | 835 (100%) | 0 (0%) |
| Malignant | 6802 (81.1%) | 6767 (99.5%) | 35 (0.5%) |

<sup>1</sup>Percentage is based on the total number of significant correlations for each group.
<sup>2</sup>Percentage of the significant correlations are based on the total number of significant correlations in the discovery cohort for each group.

**Figure 1.**
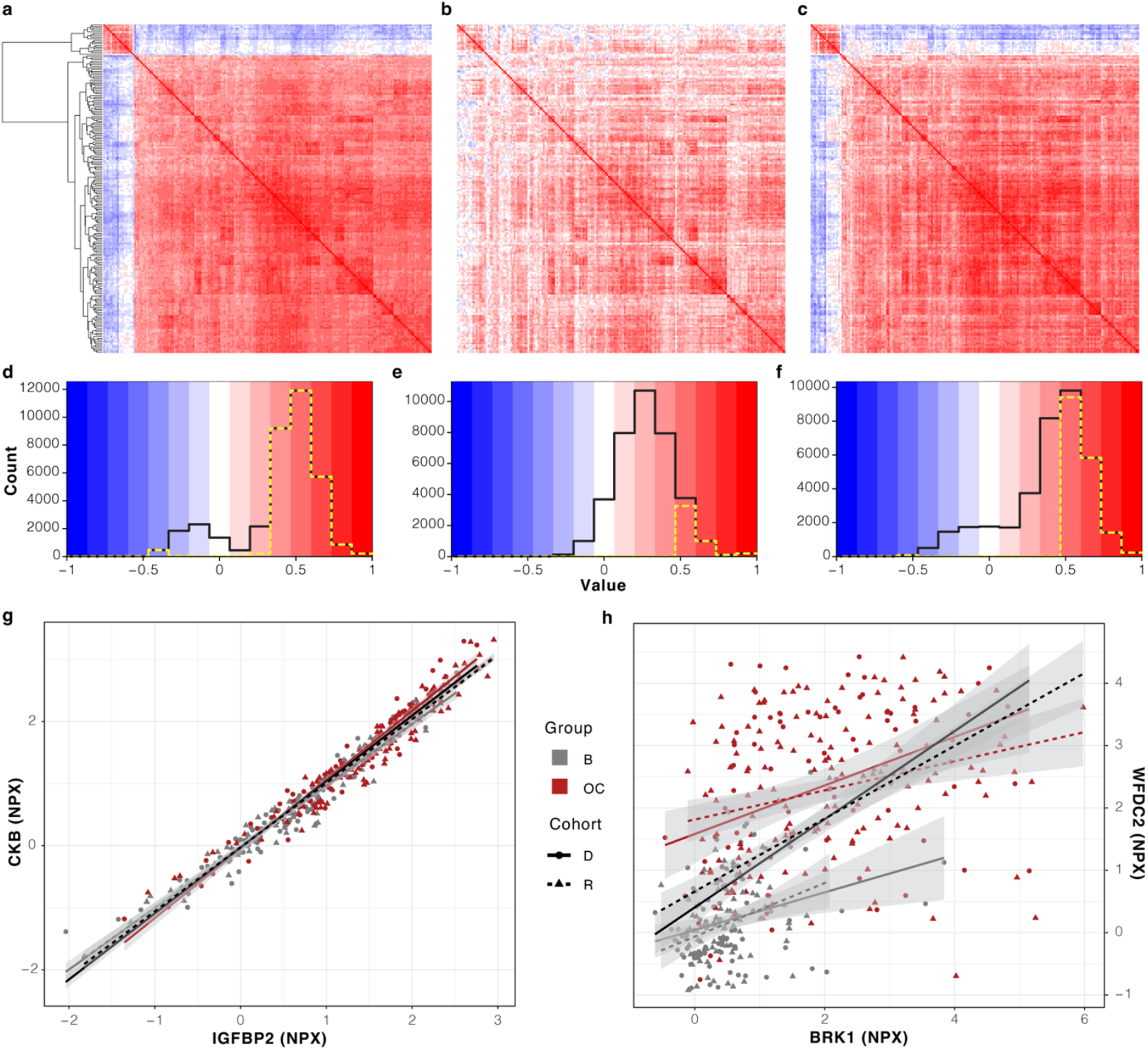
Correlation heatmaps and their corresponding correlation distributions for the discovery cohort. **a)** The protein correlations for the previously reported 191 biomarker candidates. Row-order determined unsupervised hierarchical clustering of the correlations with corresponding dendrogram to the left. **b)** As a) but for the benign group with row-order as from a). **c)** As b) for the malignant group. **d)** Histogram showing the distribution of all correlation coefficients (solid, black) from a) and the distribution of significant correlations (dashed, yellow line). **e)** As d) but with correlations coefficients from b). **f)** As d) with correlations coefficients from c). **g)** Scatterplot of the normalized protein expression (NPX) values for the protein pair CKB (y-axis) and IGFBP2 (x-axis). The ovarian cancer cases (OC) are in red, and the benign cases (B) are grey, with a linear regression line for each group in the same color. The discovery cohort (D) samples are represented by circles with a solid regression line, and the replication cohort (R) samples as triangles with a dashed regression line. The black regression lines represent the linear regression of all samples for each cohort. The shaded area around each line shows the 95% confidence interval of the regression lines. **h)** as g) but for the protein pair WFDC2 (y-axis, right) and BRK1 (x-axis).

Next, we repeated the analysis in the replication cohort where we identified the same correlation pattern of the two primary clusters as in the discovery cohort (Supplementary Figure 1a-f). The number of significant correlations and their distribution between positive and negative correlations for the replication cohort can be seen in Supplementary Table 1. In addition to the general patterns, we next evaluated if specific significant protein-protein correlations detected in the discovery cohort were also significant in the replication cohort, the summary of which can be seen in Table 1. In the combined analysis, 91.5% of the specific correlations were validated in the replication cohort (13020 / 14226) after adjustment for multiple hypothesis testing (Holm’s method), out of which 98.3% were positive and 1.7% were negative. In the separate analyses of cases and controls, 38.2% (835 / 2188) and 81.1% (6802 / 8391) of the correlations could be validated in the replication cohort after adjustment for multiple hypothesis testing (Holm’s method), respectively. For the controls, all replicated correlations were positive, while for the cases, 99.5% of replicated correlations were positive and 0.5% negative. For all validations, we saw no difference in the replication fractions stratified on positive or negative correlations (all p > 0.2, Fishers’ Exact test). The protein pair with the highest significant correlation in all groups for both cohorts was creatine kinase B-type (CKB) and insulin like growth factor binding protein 2 (IGFBP2) (Figure 1g). For that pair, the correlation-coefficients were 0.98 in the combined analyses in both the discovery and replication cohort. The protein pair with the highest significant correlation which was only found in the combined group, but not for the controls or cases separately, for both the discovery and replication cohort was WFDC2 and BRICK1 subunit of SCAR/WAVE actin nucleating complex (BRK1) (Figure 1h) with a correlation of 0.66 and 0.60 in the discovery and replication cohort, respectively.

### Ovarian cancer protein biomarker candidates show high levels of interaction

To investigate the known interactions between the 191 biomarker candidates, a protein-protein interaction network was created based on interactions listed in the STRING database^18^. Using UniProt IDs and protein names, 99.5% of the 191 proteins (190/191) were matched to the STRING database, and these 190 were subsequently used to construct the interaction network (Methods, Figure 2a). The only protein which was not found was LINE1 retrotransposable element 1 (L1RE1).

**Figure 2.**
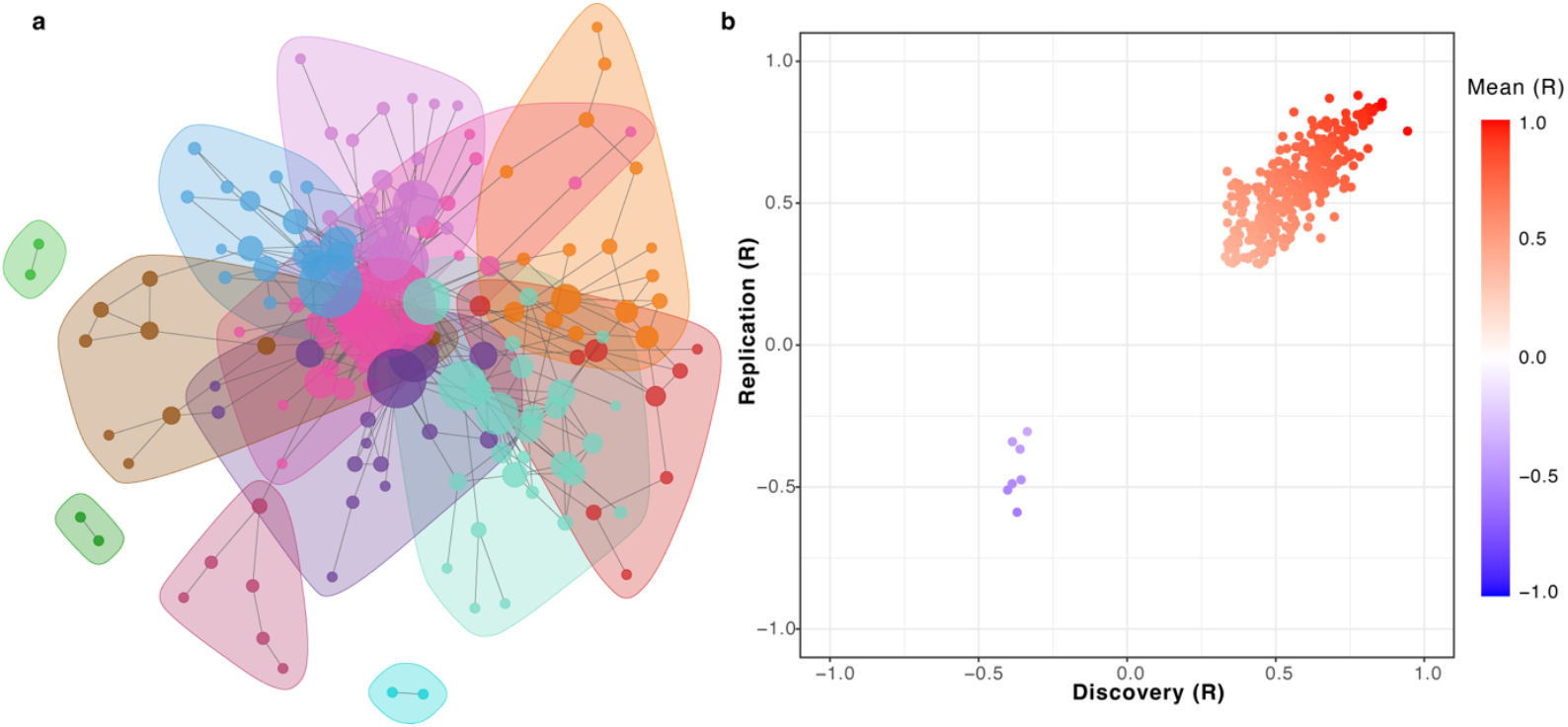
STRING protein interactions. **a)** Protein-protein interactions from the STRING database for the previously reported 191 biomarker candidates. The nodes are scaled by the number of connections, and separate clusters are illustrated by different colors. **b)** Scatterplot showing the correlations of protein pairs in a) with significant correlations for the combined benign and malignant data with the discovery coefficients on the x-axis and the replication coefficients on the y-axis.

Using an interaction score threshold of 400, 154 proteins (80.6%) was connected to at least 1 other protein. Overall, 383 interactions were found between the 190 proteins. This is significantly more protein-protein interactions (PPI, PPI enrichment p-value < 1.0 x 10^-16^) than the number of interactions expected by chance. The weighted network created from these interactions (Methods) consisted of 12 clusters out of which 9 contained 6 or more proteins. Across the whole network, we then identified the so-called hub proteins, i.e. the proteins with the most connections, or interactions, with other proteins. Here, the hub proteins with the top 3 connection counts were interleukin 6 (IL6), mucin 1 (MUC1), C-X-C motif chemokine ligand 8 (CXCL8 or IL8), and hypoxia inducible factor 1 subunit alpha (HIF1A) with 38, 23, 21 and 21 connections, respectively. The full list of interactions is available in Supplementary Data 2. A total of 315 (83.6%) protein pairs with present interactions in the STRING database also had a significant correlation in the combined control and case group for both the discovery and replication cohort (Figure 2b), with mean absolute correlation coefficients of 0.56 and 0.55 in the discovery and replication cohorts, respectively.

### Differential correlation reflects expression differences in plasma proteins between malignant and benign cases but is not replicated in tumor gene expression

Based on the large correlation differences observed, we next performed a formal differential correlation analysis (DCA) to identify specific proteins-pairs with statistically significant differences in correlation between the controls and cases. Here, we included the full protein data (N=5414) for an as complete analysis as possible. The result from the DCA was then visualized as a network (Figure 3a), filtered to include only protein pairs with significant (*q* < 0.05, adjusted for multiple hypothesis testing with Holm’s method) correlation differences (ΔR) above 0.5 between cases and controls (Supplementary Data 3). The resulting protein network contained a total of 31 proteins divided into 6 distinct clusters. The hub proteins with the top 3 connection counts were RNA polymerase-associated protein LEO1 (LEO1), kinesin family member C1 (KIFC1), and PC4 and SRSF1 interacting protein 1 (PSIP1) with 16, 6, and 4 connections respectively. Out of all proteins in the network based on the discovery cohort, 64.5% (20/31) overlapped with the 191 previously reported biomarker candidates^*5*^. Overall, we identified a high similarity between protein-protein ΔRs in the discovery cohort and the replication cohort across both positive and negative differences (R^2^ = 0.83, Methods, Figure 3b). In addition to the overall similar patterns, 67.7% (21/31) of the individual connections detected in the discovery cohort remained significant in the replication cohort (*q* < 0.05, adjusted for multiple hypothesis testing with Holm’s method).The protein pair with the highest differential correlation in the discovery cohort, and a significant ΔR > 0.5 for both the discovery and replication cohort was LEO1 and RNA binding fox-1 homolog 3 (RBFOX3) (Figure 3c), with a ΔR of 0.86 and 0.62 for discovery and replication respectively. For this pair, the correlation coefficients in the discovery cohort were - 0.01 for the controls and 0.85 for the cases. In the replication cohort, the correlation coefficients for the controls and cases were 0.17 and 0.79, respectively.

**Figure 3.**
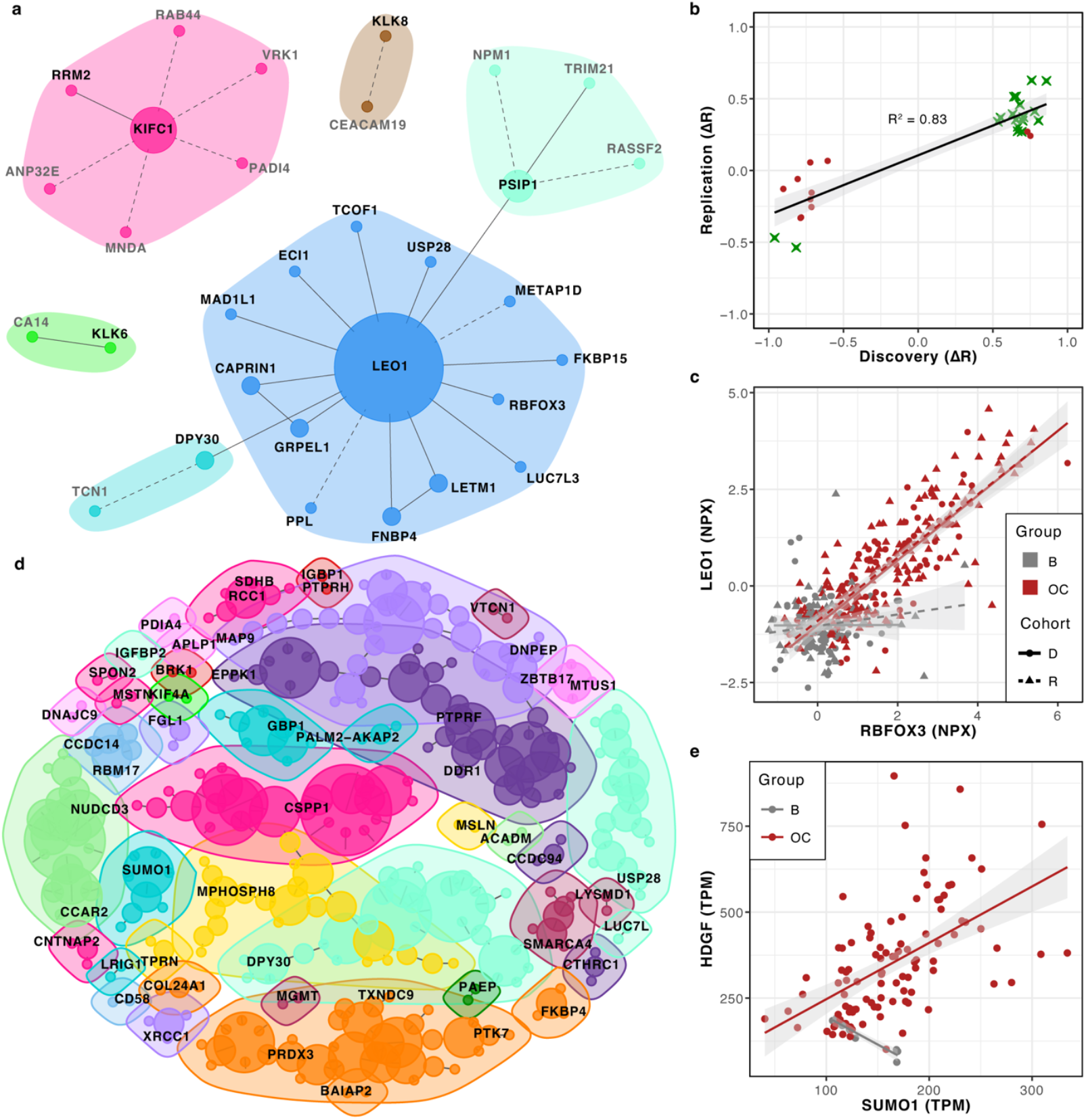
Differential correlation analysis (DCA) networks. **a)** Network of the differentially correlated plasma proteins between the malignant and benign group for proteins with a significant (adjusted p < 0.05) correlation difference (ΔR) above 0.5 in the discovery cohort. Dashed edges illustrate correlations which were not significant in the replication cohort. Black labels correspond to proteins among the previously reported 191 biomarker candidates. Node sizes are scaled by the number of connections. **b)** Scatterplot of the correlation differences for the discovery (x-axis) and replication (y-axis) cohort. Green stars represent protein pairs with a significant differential correlation in both discovery and replication, and red circles represent protein pairs with non-significant differential correlations in the replication cohort. **c)** Scatterplot of the normalized protein expression (NPX) values of the pair LEO1 (y-axis) and RBFOX3 (x-axis). The cases (OC) are in red, and the controls (B) are grey, each group with a linear regression line in the same color. The discovery cohort (D) samples are circles with a solid regression line, and the replication cohort (R) samples are triangles with a dashed regression line. The shaded area around each line represents the 95% confidence interval. **d)** Network of the differentially correlating tumor RNA for the replication cohort between malignant and benign cases for the gene-equivalents to the protein panel. The network is filtered by significance (adjusted p < 0.5), a correlation difference (ΔR) above 0.5, and for clusters which contain at least 1 protein among the 191 previously reported biomarker candidates (labelled in black). Nodes are scaled by the number of connections. **e)** Scatterplot of RNA expression (TPM) for the transcript pair HDGF (y-axis) and SUMO1 (x-axis). The malignant cases are in red, and the benign cases are gray, each group with a linear regression line in the same color. The shaded area around each line represents the 95% confidence interval.

Next, we performed a second DCA analysis in the replication cohort based on tumor gene expression (TPM, Methods). A total of 5424 transcripts were found to overlap with the 5416 proteins from the proteomics panel, with 4878 transcripts corresponding to 4876 proteins remaining after quality control (Methods). Using these 4878 transcripts, the differential correlations were calculated, and a network was constructed using the significant correlations (*q* < 0.05, adjusted for multiple hypothesis testing with Holm’s method) with a correlation difference (ΔR) over 0.5. The final network consisted of 759 differentially correlated transcript pairs, with a total of 952 individual transcripts (Supplementary Figure 2, Supplementary Data 4). As above, hub transcripts were identified and the top hub transcripts with the highest connection counts encode the proteins NRAS proto-oncogene, GTPase (NRAS), ENY2 transcription and export complex 2 subunit (ENY2), glia maturation factor gamma (GMFG), glyoxalase I (GLO1), tubulin tyrosine ligase like 4 (TTLL4), and, estrogen receptor binding site associated antigen 9 (EBAG9) with 9, 8, 7, 7, 7 and 7 connections, respectively. A focused subnetwork including only clusters with at least 1 transcript encoding one of 191 previously identified plasma protein biomarkers can be seen in Figure 3d. This network included 27.2% (52/191) of the transcripts, corresponding to 5.5% of the transcripts of the full network (supplementary data). An example of a protein pair in the subnetwork where one of the transcripts encode one of the 191 proteins is heparin binding growth factor (HDGF) and small ubiquitin like modifier 1 (SUMO1) (Figure 3e), where SUMO1 was amongst the 191 protein biomarker candidates and HDGF was not. Lastly, we investigated if any of the identified differentially correlated pairs overlapped between the resulting full RNA network (Supplementary Figure 2) and the full protein network (Figure 3a) networks and found no overlapping connections.

## Discussion

In this study we identified large differences in correlation patterns between differentially expressed proteins in plasma from women with either benign or malignant ovarian tumors and in tumor gene expression from the same women. Based on analyses in two separate cohorts, the majority of results for plasma proteins identified in one cohort were replicated in the second. However, the identified differences were not the same for the analyses carried out in proteins as compared to analyses carried out based on RNA expression in matched individuals. This is in line with our previously published results^12^ comparing simple correlations between circulating plasma proteins and tumor gene expression. Identified differences in co-expression between benign and malignant tumors not generally being reflected in the plasma proteome, and vice-versa, also suggests an important role of the tumor microenvironment in the overall interpretation of plasma proteomics in relation to specific diseases^21^. These observations do not however limit the potential use of identified case-control differences in plasma proteins as biomarkers for a specific disease but instead emphasize the need for carefully constructed future studies to identify causal relationships for the discovery of new drug targets, or repurposing of existing drugs^22^. We also identified large networks of co-expressed proteins based on plasma abundance levels which were largely driven by patterns seen specifically in patients diagnosed with malignant tumors and to a lesser degree in patients diagnosed with benign tumors, both in relation to distributions of correlation coefficients and in the fraction of these that were significant. The majority of the protein-pairs identified here with high circulating concentrations in plasma have previously reported interactions in the STRING database. When restricted to the 191 previously suggested plasma protein biomarkers for ovarian cancer, the STRING-interaction network showed a highly interconnected network with several well-connected proteins. This network has significantly more interactions than expected by chance alone, further supporting an underlying co-regulatory mechanism between several of the potential biomarker candidates^12^. It should be noted however that the interaction score from the STRING database does not necessarily represent biological co-regulation and that the database is not specific to cancer or tumor development. The vast majority (> 80%), however, of the STRING-connections identified here were also shown to have significant correlations in measured plasma proteomics solidifying the observed interactions. In addition, the proteins with the highest interaction counts (hub-proteins) in the focused network are all highly implicated with ovarian cancer relevance in the existing literature. The top connected proteins were IL6, MUC1, CXCL8, and HIF1. IL6 is a cytokine with involvement in immune responses and inflammatory processes, and its overexpression has been connected to cancer and other diseases^23,24^. It has previously been shown to be of prognostic value for ovarian cancer^*25*^, where high serum levels of IL6 in ovarian cancer patients has been connected to increased risk of relapse, lower overall survival, and to drug resistance against cancer therapeutics^*25–27*^. MUC1, a membrane-bound protein involved in cell-signaling, has previously been reported to be overexpressed in the majority of epithelial ovarian cancer and is connected to cancer progression^*28*^. HIF1A has been shown to have a lower level of expression in OC compared to benign tumors, both when looking at tumor transcript levels^*29*^ and plasma protein levels^*5*^. CXCL8 is a proinflammatory CXC chemokine known to be involved in several signaling pathways in many cancer types, including ovarian cancer, where it promotes proliferation and metastasis^30,31^. CXCL8 has also been suggested as both a predictive biomarker and a therapeutic target for cervical cancer^32,33^. IL6, HIF1A, and CXCL8 are also all involved in angiogenesis^*30,34,35*^, and both IL6 and CXCL8 have previously also been suggested as potential plasma biomarkers for ovarian cancer^36,37^.

When comparing differential correlation between benign and malignant tumors using our full plasma proteomics data and comparing that with the same analysis using RNA expression levels in tissue, we observed a higher number of significantly different co-regulated pairs on the RNA-level as compared to the protein level. Interestingly, no overlap was found between the differentially correlated pairs in the respective data type. The latter further supports the importance of studying also the tumor microenvironment to fully understand the relationship between circulating plasma biomarkers and the tumors themselves. The two transcripts with the highest connection counts (hub transcripts) in the generated networks were *NRAS* and *ENY2*. Both transcripts have previously been associated with tumor development in general, and in relation to ovarian cancer, specifically. Mutations in *NRAS* have been associated with tumor development in ovarian cancer, with high mutation frequencies in low-grade serous ovarian cancer^38^ and to a lesser degree also in high-grade serous ovarian cancers^39^. *ENY2* has been suggested as a pan-cancer marker based on tissue expression levels^40^, and has specifically been noted to have increased copy-numbers in ovarian cancers with a resulting increased expression ^41^. Both proteins were included in the assay used in our previous study^12^ but were not brought forward as potential plasma biomarkers for ovarian cancer. We found no difference in NRAS levels of circulating plasma (unadjusted p-values > 0.42 in both discovery and replication) between women with benign and malignant ovarian tumors. For ENY2, the differences detected in plasma were nominally significant in both our discovery and replication cohort (unadjusted p-value < 3.8 x 10^-5^) but did not remain significant after adjustment for multiple hypothesis testing. These examples illustrate the difficulties in interpretating function in relation to tumor biology when examining plasma proteomics alone.

Our study is limited by several factors. The cohorts used are both Swedish and it is known that several factors, including lifestyle, genetics, and ethnicity, can have strong effects on the circulating plasma proteome^42^ which could limit the generalization of specific protein-protein correlations detected here. In addition, although the assay (PEA) used for characterization of the plasma proteome is highly sensitive^43^ there is a lower limit of detection (LOD) for each analyte. In our analyses of correlation separately in cases and controls, we noticed a considerably lower number of significant correlations in the controls compared to the cases, even though the sample sizes were similar (85 vs 86). This could be due to biological reasons as a higher number of proteins are upregulated in malignant cases. Alternatively, the measurable range of the assays, i.e. signal to noise ratio, could be lower in the controls as compared to the cases. Possibly, a more sensitive assay would have detected even lower concentrations, which in turn could have resulted in more robust correlations between proteins in the benign group as well. Our results are strengthened by several factors. First, our study design with two geographically different cohorts, where one was used strictly as discovery and the second for validation, reduced the number of false positive results. Secondly, we characterized a large set of proteins performed total RNA-sequencing analyses in tumor tissue from the same women, allowing us to directly compare these two data types. Lastly, the majority of the detected protein-protein correlations could be replicated not only in our validation cohort, but also by analysis of previously known interactions available in the STRING-database.

In conclusion, our findings show large contrasts in plasma protein correlation patterns between women with benign or malignant ovarian tumors, highlighting the complex biological differences between these conditions. While no direct connections between differential expression of the tumor RNA and plasma proteome were identified, these findings suggest that tumor-induced changes might affect downstream processes. This highlights the potential in studying tumor-host interactions and the tumor microenvironment. The identified protein interactions and correlations may provide a new perspective on disease mechanisms and help guide the discovery of biomarkers or potential drug targets.

## Data Availability

The raw data used is available at Science for Life Laboratories (SciLifeLab) Data Repositories with the following accession numbers: https://doi.org/10.17044/scilifelab.27233412 (protein-data) and https://doi.org/10.17044/scilifelab.28342484 (RNA-data.

https://doi.org/10.17044/scilifelab.28342484

## Acknowledgements

This study was funded by; the Swedish Research Council 2022-00857 (SE), the Swedish Cancer Foundation 220604FE (SE), the Swedish Cancer Foundation 232874PJ (SE) and the Swedish Cancer Foundation 190008PJ (UG). The computations and data handling were enabled by resources in project sens2023504 provided by the National Academic Infrastructure for Supercomputing in Sweden (NAISS) at Uppsala Multidisciplinary Center for Advanced Computational Science (UPPMAX) partially funded by the Swedish Research Council through grant agreement no. 2022-06725. Additional support was provided by Paul-Theodor Pyl from the Swedish Bioinformatics Advisory Program at the National Bioinformatics Infrastructure Sweden (NBIS).

## Author contributions

**Mikaela Moskov:** Conceptualization, Methodology, Software, Validation, Formal analysis, Investigation, Writing - Original Draft, Writing - Review & Editing, Visualization **Julia Hedlund Lindberg:** Investigation, Writing - Review & Editing **Ulf Gyllensten:** Resources, Funding Acquisition, Writing - Review & Editing **Stefan Enroth:** Conceptualization, Methodology, Resources, Writing - Original Draft, Writing - Review & Editing, Supervision, Project administration, Funding acquisition

## Data availability

Raw data is located in controlled access data storage at the Swedish Science for Life Laboratories (SciLifeLab) Data Repositories with the following accession numbers: https://doi.org/10.17044/scilifelab.27233412 (protein-data) and https://doi.org/10.17044/scilifelab.28342484 (RNA-data). Results data underlying the figures in this publication can be found in the Supplementary Data.

## List of Supplementary Materials

### Supplementary Figures and Tables

**Supplementary Figure 1**. Correlation heatmaps and distribution for the replication cohort.

**Supplementary Figure 2**. Differential correlation network for the RNA of the replication cohort

**Supplementary Table 1**. Significant correlations in the replication cohort and the distribution between positive and negative correlations.

### Supplementary Data

**Supplementary Data 1**. Correlation data for Figure 1a-f and Supplementary Figure 1a-f

**Supplementary Data 2**. Protein pairs and interaction scores of the STRING data used in Figure 2a

**Supplementary Data 3**. Differential correlation data for the protein network in Figure 3ab

**Supplementary Data 4**. Differential correlation data for the RNA network in Figure 3d and Supplementary Figure 2

## Abbreviations

BRK1: BRICK1 subunit of SCAR/WAVE actin nucleating complex
CA-125: Cancer antigen 125
CKB: Creatine kinase B-type
CXCL8: C-X-C motif chemokine ligand 8, IL8
DCA: Differential correlation analysis
EBAG9: Estrogen receptor binding site associated antigen 9
ENY2: GTPase ENY2 transcription and export complex 2 subunit
GLO1: Glyoxalase I
GMFG: Glia maturation factor gamma
HDGF: Heparin binding growth factor
HIF1A: Hypoxia inducible factor 1 subunit alpha
IGFBP2: Insulin like growth factor binding protein 2
IL6: Interleukin 6
KIFC1: Kinesin family member C1
LEO1: RNA polymerase-associated protein LEO1
LOD: Limit of detection
MUC1: Mucin-1
MUC16: Mucin-16
NPX: Normalized protein expression
NRAS: NRAS proto-oncogene
PEA: Proximity extension assay
PSIP1: PC4 and SRSF1 interacting protein 1
RBFOX3: RNA binding fox-1 homolog 3
SUMO1: Small ubiquitin like modifier 1
TPM: Transcripts per million
TTLL4: Tubulin tyrosine ligase like 4
WFDC2: Human epididymis protein 4, HE4

## Notes

### Competing Interest Statement

The authors have declared no competing interest.

### Author Declarations

The Swedish Ethical Review Authority (2023-05515-02, extension of Regional Ethics Committee in Uppsala Dnr:2016/145) and Goteborg (Dnr: 201-15) gave ethical approval for this work.

